## Supplemental tables and figures for "Non-random Mating Patterns in Education, Mental, and Somatic Health: A Population Study on Within- and Cross-Trait Associations"

**Table of contents**

**Supplemental information for prospective analyses**
Supplemental Figure S1. Within person correlations S2

**Supplemental information for prospective analyses with odds ratios**
Supplemental Figure S2. Within person associations S3
Supplemental Figure S3. Within and across-trait partner associations S4
Supplemental Table S1. Results from logistic regression among partners and in-laws S5

**Supplemental information for cross-sectional analyses**
Supplemental Table S2. Prevalence rates S6
Supplemental Table S3. Correlations between relatives S7
Supplemental Figure S4. Prevalence rates among affected and unaffected partners S8
Supplemental Figure S5. Correlations between partners with different adjustments S9
Supplemental Figure S6. Within person correlations S10
Supplemental Figure S7. Partner correlations adjusted for age S11
Supplemental Figure S8. Partner correlations adjusted for age and GPA S12
Supplemental Figure S9. Partner correlations adjusted for age and EA S13

**Supplemental information for methods**
Supplemental Script S1. Calculation of the in-law inflation factor (IIF) S14
Supplemental Script S2. Potential bias in tetrachoric correlations for skewed variables S15
Supplemental Script S3. Potential bias in the product of tetrachoric correlations S16

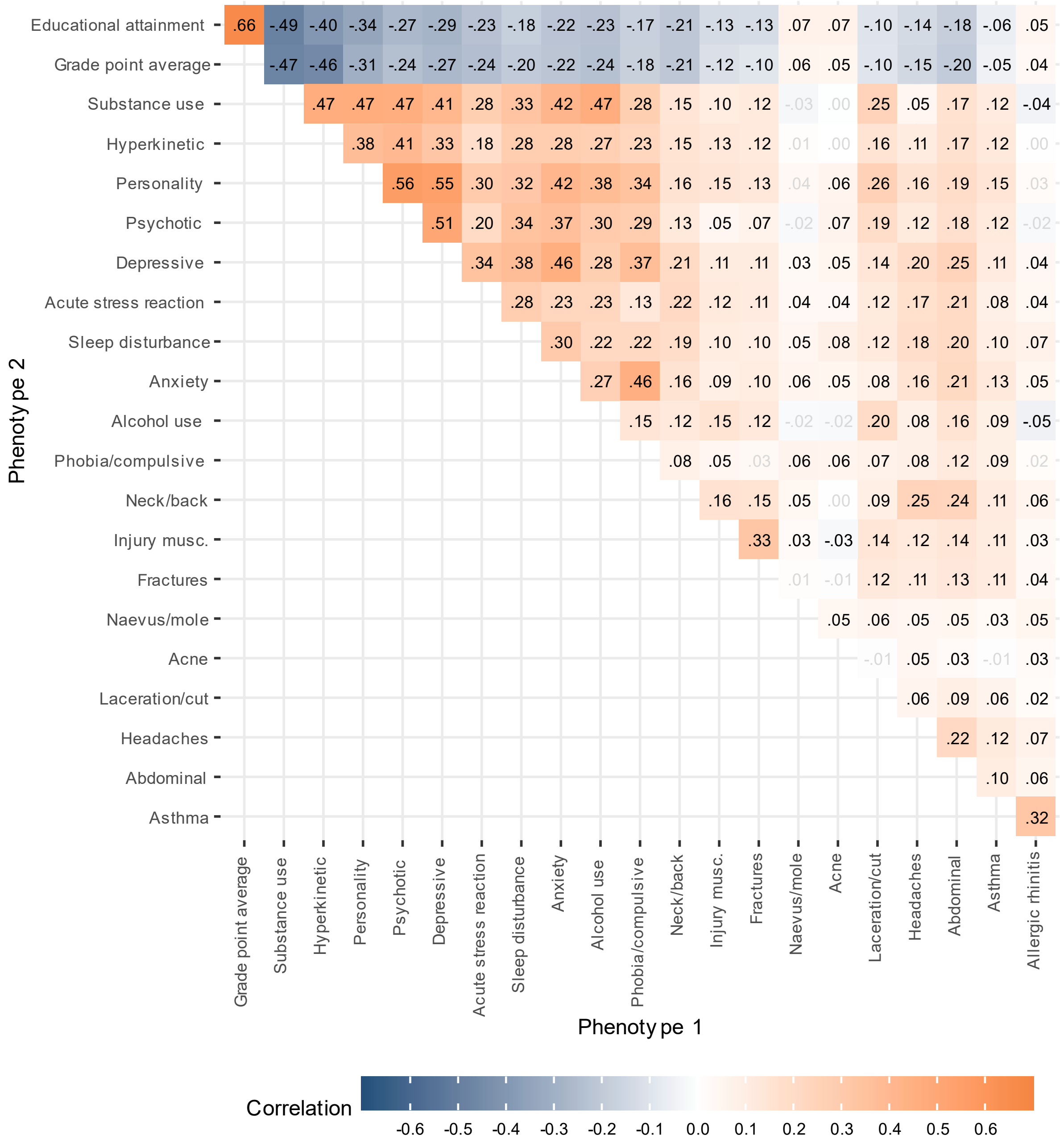


**Supplemental Figure S1.** Within person correlations for educational outcomes, 10 mental health conditions, and 10 somatic health conditions, 10 to 5 years before first child. Adjusted for age and sex. Correlations shown in black have p-values < 0.05 after adjusting for the false discovery rate.

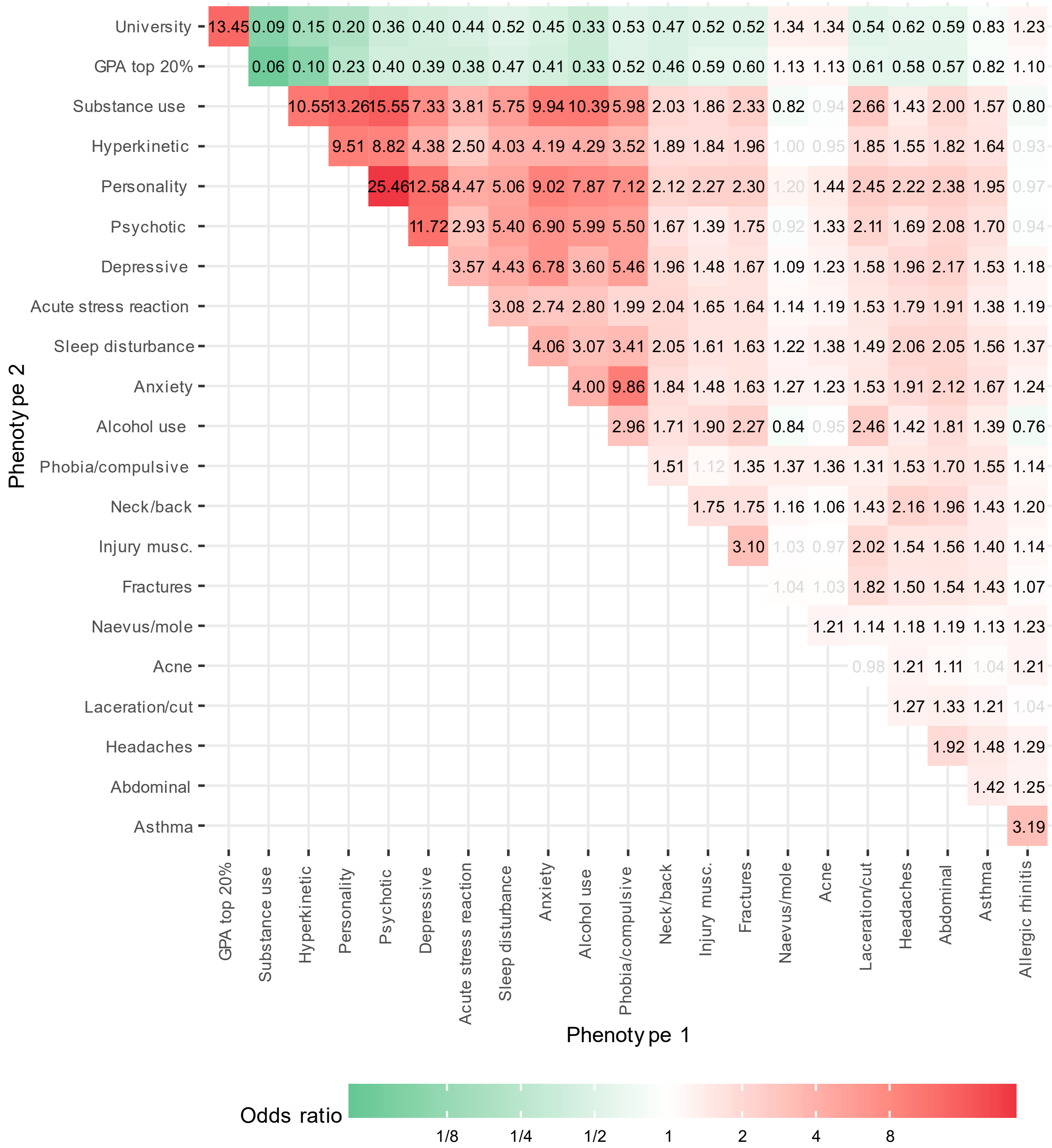


**Supplemental Figure S2.** Odds rations for within person associations between educational outcomes, 10 mental health conditions, and 10 somatic health conditions, 10 to 5 years before first child. Adjusted for age and sex. Correlations shown in black have p-values < 0.05 after adjusting for the false discovery rate.

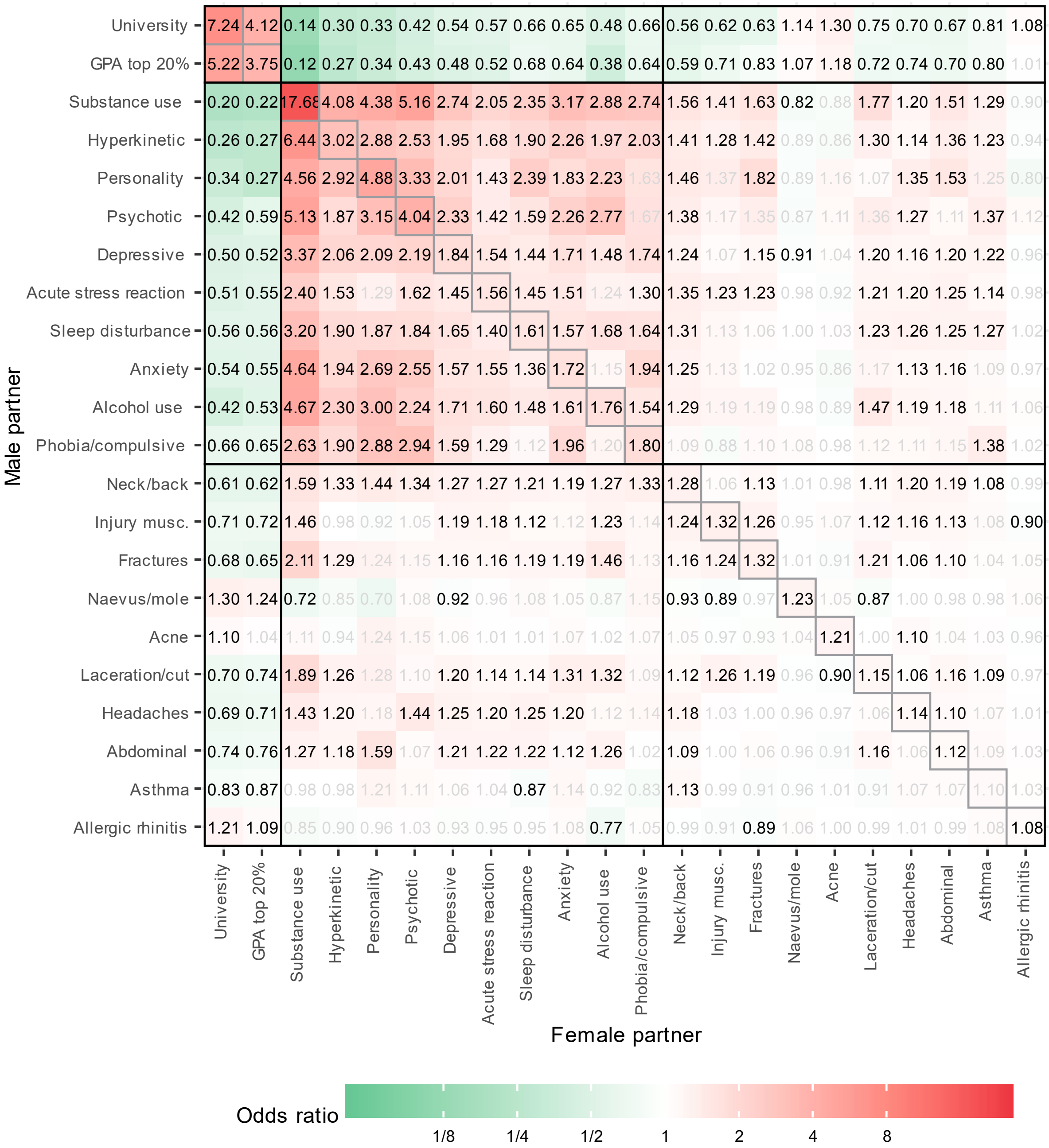


**Supplemental Figure S3**. Odds ratios for within and across-trait partner associations between educational outcomes, 10 mental health conditions, and 10 somatic health conditions, 10 to 5 years before first child, all adjusted for age. Correlations shown in black have p-values < 0.05 after adjusting for the false discovery rate.

**Supplemental Table S1.** Results from multiple binary logistic regressions of each of the educational outcomes and the 10 mental and 10 somatic health conditions on the corresponding traits in partners, siblings, and in-laws. Health conditions were measured 10 to 5 years before a couple had their first child. Results expressed as adjusted odds ratios (OR), including 95% confidence intervals. Significant adjusted associations with siblings-in-law indicate deviations from direct assortment.

| | OR(partner) | OR(sibling) | OR(in-law) |
|---|---|---|---|
| University education | 5.35 [5.11, 5.59] | 3.58 [3.44, 3.73] | 1.76 [1.69, 1.83] |
| GPA among top 20% | 2.76 [2.55, 3.00] | 4.57 [4.22, 4.94] | 1.54 [1.42, 1.68] |
| Substance use disorders | 16.07 [12.83, 20.12] | 4.91 [3.61, 6.68] | 3.74 [2.75, 5.08] |
| Hyperkinetic disorder | 2.73 [2.17, 3.43] | 6.40 [5.37, 7.62] | 1.37 [1.03, 1.81] |
| Personality disorder | 1.99 [0.49, 8.06] | 3.23 [1.19, 8.74] | 3.39 [1.25, 9.18] |
| Psychotic disorders | 2.91 [1.53, 5.51] | 3.41 [2.08, 5.59] | 0.96 [0.39, 2.32] |
| Depressive disorder | 1.77 [1.62, 1.94] | 2.36 [2.17, 2.56] | 1.27 [1.15, 1.39] |
| Acute stress reaction | 1.46 [1.31, 1.63] | 2.72 [2.48, 2.98] | 1.13 [1.01, 1.26] |
| Sleep disturbance | 1.58 [1.38, 1.82] | 1.89 [1.66, 2.15] | 1.05 [0.90, 1.24] |
| Anxiety disorder | 1.59 [1.29, 1.96] | 2.68 [2.28, 3.16] | 1.18 [0.94, 1.47] |
| Alcohol use disorders | 1.69 [1.16, 2.46] | 2.04 [1.39, 2.98] | 1.41 [0.91, 2.19] |
| Phobia/compulsive disorder | 1.73 [1.10, 2.71] | 2.17 [1.49, 3.15] | 1.38 [0.85, 2.25] |
| Neck/back symptom | 1.25 [1.19, 1.31] | 1.49 [1.42, 1.57] | 1.14 [1.08, 1.20] |
| Injury musculoskeletal | 1.29 [1.17, 1.41] | 1.63 [1.49, 1.79] | 1.13 [1.03, 1.25] |
| Fractures | 1.27 [1.14, 1.42] | 1.42 [1.27, 1.58] | 1.19 [1.06, 1.33] |
| Naevus/mole | 1.18 [1.09, 1.27] | 1.61 [1.50, 1.73] | 1.06 [0.98, 1.14] |
| Acne | 1.14 [1.00, 1.30] | 2.33 [2.10, 2.59] | 0.96 [0.83, 1.11] |
| Laceration/cut | 1.18 [1.08, 1.28] | 1.21 [1.11, 1.32] | 1.04 [0.96, 1.14] |
| Headaches | 1.11 [1.03, 1.19] | 1.49 [1.39, 1.59] | 1.05 [0.98, 1.12] |
| Abdominal pain | 1.11 [1.04, 1.18] | 1.41 [1.33, 1.50] | 1.06 [1.00, 1.13] |
| Asthma | 1.12 [0.99, 1.27] | 2.50 [2.28, 2.75] | 1.04 [0.92, 1.18] |
| Allergic rhinitis | 1.09 [1.00, 1.18] | 2.03 [1.89, 2.18] | 1.02 [0.94, 1.12] |

**Supplemental Table S2.** List of ICPC-2 codes for the mental and somatic health condition, prevalence in the sample (including education), and prevalence among partners and relatives of affected individuals, measured cross sectionally in 2015-2019.

| | | Index | | Partner of affected | | Sibling of affected | | In-law of affected | |
|---|---|---|---|---|---|---|---|---|---|
| Variable | ICPC-2 codes | n | % | n | % | n | % | n | % |
| University education | | 93,303 | 49.84 | 64,110 | 68.89 | 48,682 | 61.23 | 46,001 | 58.16 |
| GPA among top 20% | | 26,824 | 20.15 | 8,058 | 37.43 | 7,654 | 42.38 | 5,085 | 32.38 |
| Substance use disorders | P18, P19 | 2,899 | 1.54 | 724 | 24.97 | 163 | 8.22 | 143 | 6.76 |
| Hyperkinetic disorder | P81 | 5,216 | 2.78 | 618 | 11.85 | 542 | 14.03 | 223 | 5.60 |
| Personality disorder | P80 | 1,652 | 0.88 | 64 | 3.87 | 53 | 4.39 | 25 | 1.99 |
| Psychotic disorders | P72, P98, P73 | 1,965 | 1.05 | 80 | 4.07 | 95 | 6.26 | 29 | 1.88 |
| Depressive disorder | P76 | 21,811 | 11.61 | 4,126 | 18.92 | 2,966 | 17.59 | 2,352 | 13.47 |
| Acute stress reaction | P02 | 24,996 | 13.30 | 5,442 | 21.77 | 3,320 | 16.81 | 2,564 | 12.67 |
| Sleep disturbance | P06 | 16,807 | 8.94 | 2,424 | 14.42 | 1,619 | 12.17 | 1,312 | 9.63 |
| Anxiety disorder | P74 | 8,202 | 4.36 | 648 | 7.90 | 560 | 8.83 | 353 | 5.34 |
| Alcohol use disorders | P15, P16 | 2,505 | 1.33 | 158 | 6.31 | 91 | 4.83 | 64 | 3.29 |
| Phobia/compulsive disorder | P79 | 3,702 | 1.97 | 146 | 3.94 | 128 | 4.44 | 93 | 3.12 |
| Neck/back symptom/complaint | L01, L02, L03 | 44,851 | 23.87 | 12,446 | 27.75 | 8,342 | 23.27 | 7,633 | 20.90 |
| Injury musculoskeletal | L81 | 12,271 | 6.53 | 1,094 | 8.92 | 911 | 9.16 | 800 | 7.96 |
| Fractures | L72, L73, L74, L75, L76 | 10,641 | 5.66 | 764 | 7.18 | 694 | 8.01 | 556 | 6.40 |
| Naevus/mole | S82 | 29,454 | 15.67 | 5,030 | 17.08 | 4,535 | 18.26 | 3,614 | 14.47 |
| Acne | S96 | 7,721 | 4.11 | 326 | 4.22 | 638 | 10.00 | 356 | 5.53 |
| Laceration/cut | S18 | 15,377 | 8.18 | 1,388 | 9.03 | 1,299 | 10.15 | 1,152 | 9.11 |
| Headaches | N89, N90, N01, N95 | 32,011 | 17.03 | 5,148 | 16.08 | 4,808 | 18.59 | 4,136 | 15.64 |
| Abdominal pain/cramps general | D01 | 36,242 | 19.29 | 6,878 | 18.98 | 5,427 | 18.73 | 4,809 | 16.17 |
| Asthma | R96 | 11,415 | 6.07 | 800 | 7.01 | 1,069 | 11.51 | 598 | 6.35 |
| Allergic rhinitis | R97 | 23,335 | 12.42 | 3,224 | 13.82 | 3,447 | 17.81 | 2,222 | 11.38 |

**Supplemental Table S3.** Correlations between relatives in educational outcomes and 10 mental and 10 somatic health conditions measured cross-sectionally in 2015-2019, including 95% confidence intervals, along with tests of deviations from direct assortment. Adjusted for sex and year of birth.

| Variable | r(partner) | r(sibling) | r(inlaw) | Inlaw inflation factor (IIF)* | Deviation from direct assortment, p-value** |
|---|---|---|---|---|---|
| Educational attainment | 0.48 [0.47, 0.48] | 0.40 [0.40, 0.40] | 0.29 [0.28, 0.29] | 1.50 | <1.00e-99 |
| Grade point average | 0.42 [0.42, 0.43] | 0.52 [0.52, 0.53] | 0.29 [0.29, 0.30] | 1.33 | <1.00e-99 |
| Substance use disorders | 0.67 [0.65, 0.70] | 0.32 [0.29, 0.36] | 0.27 [0.24, 0.31] | 1.25 | 0.006 |
| Hyperkinetic disorder | 0.33 [0.30, 0.36] | 0.38 [0.36, 0.40] | 0.13 [0.10, 0.16] | 1.03 | 0.805 |
| Personality disorder | 0.24 [0.17, 0.30] | 0.24 [0.19, 0.29] | 0.10 [0.05, 0.16] | 1.83 | 0.234 |
| Psychotic disorders | 0.25 [0.19, 0.31] | 0.23 [0.19, 0.27] | 0.02 [-0.04, 0.07] | 0.31 | 0.236 |
| Depressive disorder | 0.25 [0.23, 0.27] | 0.17 [0.16, 0.19] | 0.07 [0.05, 0.08] | 1.57 | 0.002 |
| Acute stress reaction | 0.25 [0.23, 0.26] | 0.19 [0.18, 0.20] | 0.06 [0.05, 0.08] | 1.36 | 0.023 |
| Sleep disturbance | 0.17 [0.15, 0.19] | 0.13 [0.11, 0.14] | 0.05 [0.03, 0.06] | 2.26 | 0.003 |
| Anxiety disorder | 0.17 [0.14, 0.20] | 0.18 [0.16, 0.20] | 0.05 [0.02, 0.07] | 1.53 | 0.280 |
| Alcohol use disorders | 0.28 [0.23, 0.33] | 0.20 [0.15, 0.24] | 0.11 [0.07, 0.16] | 2.09 | 0.023 |
| Phobia/compulsive disorder | 0.14 [0.09, 0.18] | 0.13 [0.10, 0.17] | 0.07 [0.03, 0.11] | 3.68 | 0.026 |
| Neck/back symptom | 0.12 [0.11, 0.13] | 0.11 [0.10, 0.12] | 0.05 [0.04, 0.06] | 3.35 | 4.50e-09 |
| Injury musculoskeletal | 0.09 [0.07, 0.12] | 0.08 [0.07, 0.10] | 0.04 [0.02, 0.06] | 4.71 | 0.006 |
| Fractures | 0.07 [0.04, 0.10] | 0.08 [0.06, 0.10] | 0.02 [-0.00, 0.04] | 2.70 | 0.387 |
| Naevus/mole | 0.07 [0.06, 0.09] | 0.13 [0.11, 0.14] | 0.01 [0.00, 0.03] | 1.58 | 0.403 |
| Acne | 0.04 [0.01, 0.08] | 0.16 [0.14, 0.19] | 0.02 [-0.01, 0.05] | 2.95 | 0.355 |
| Laceration/cut | 0.06 [0.04, 0.08] | 0.08 [0.06, 0.09] | 0.04 [0.02, 0.05] | 7.79 | 0.001 |
| Headaches | 0.06 [0.04, 0.07] | 0.11 [0.10, 0.12] | 0.03 [0.01, 0.04] | 3.95 | 0.006 |
| Abdominal pain | 0.09 [0.07, 0.10] | 0.09 [0.08, 0.10] | 0.02 [0.01, 0.03] | 2.97 | 0.021 |
| Asthma | 0.04 [0.01, 0.06] | 0.19 [0.18, 0.21] | 0.02 [-0.00, 0.04] | 2.67 | 0.301 |
| Allergic rhinitis | 0.04 [0.02, 0.06] | 0.18 [0.17, 0.19] | -0.00 [-0.01, 0.01] | -0.07 | 0.330 |

Notes: * IIF is expected to equal 1.00 under direct assortment. The cross-sectional partner correlations can be influenced by convergence, generally reducing the IIF. This may cancel out effects of indirect assortment and social stratification, which generally increases IIF, thereby limiting the interpretability of IIF among established couples. ** The p-value arises from comparing a constrained model, where in-law correlation is the product of partner and sibling correlations, to an unconstrained model with independent estimates for each relationship type. A low p-value signifies a poor fit for direct assortment. The p-values are adjusted for false discovery rate using the Benjamini-Hochberg method.

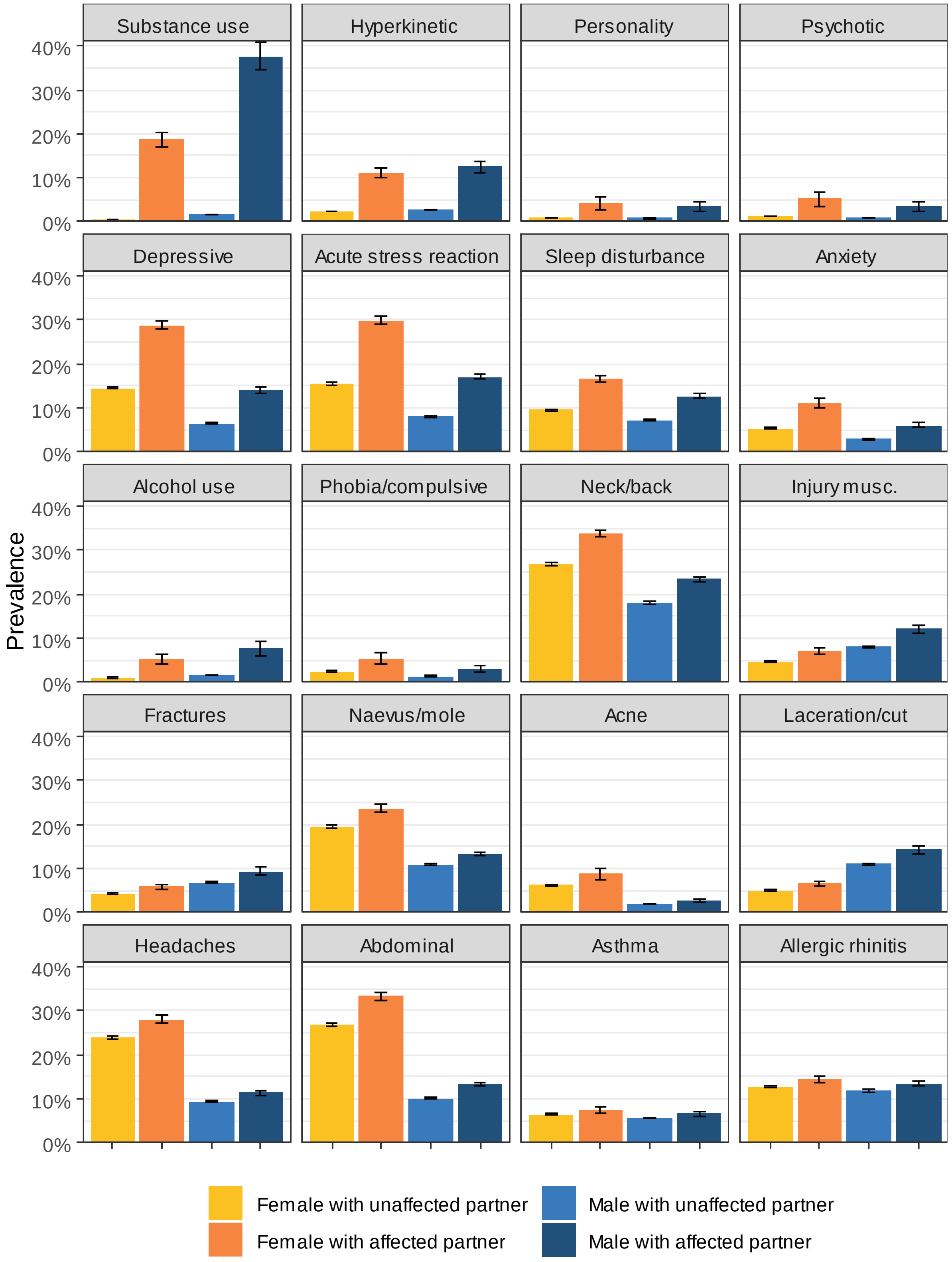


**Supplemental Figure S4**. Prevalence of 10 mental and 10 somatic health conditions among males and females with unaffected and affected partners, measured cross sectionally in 2015-2019. Including 95% confidence intervals.

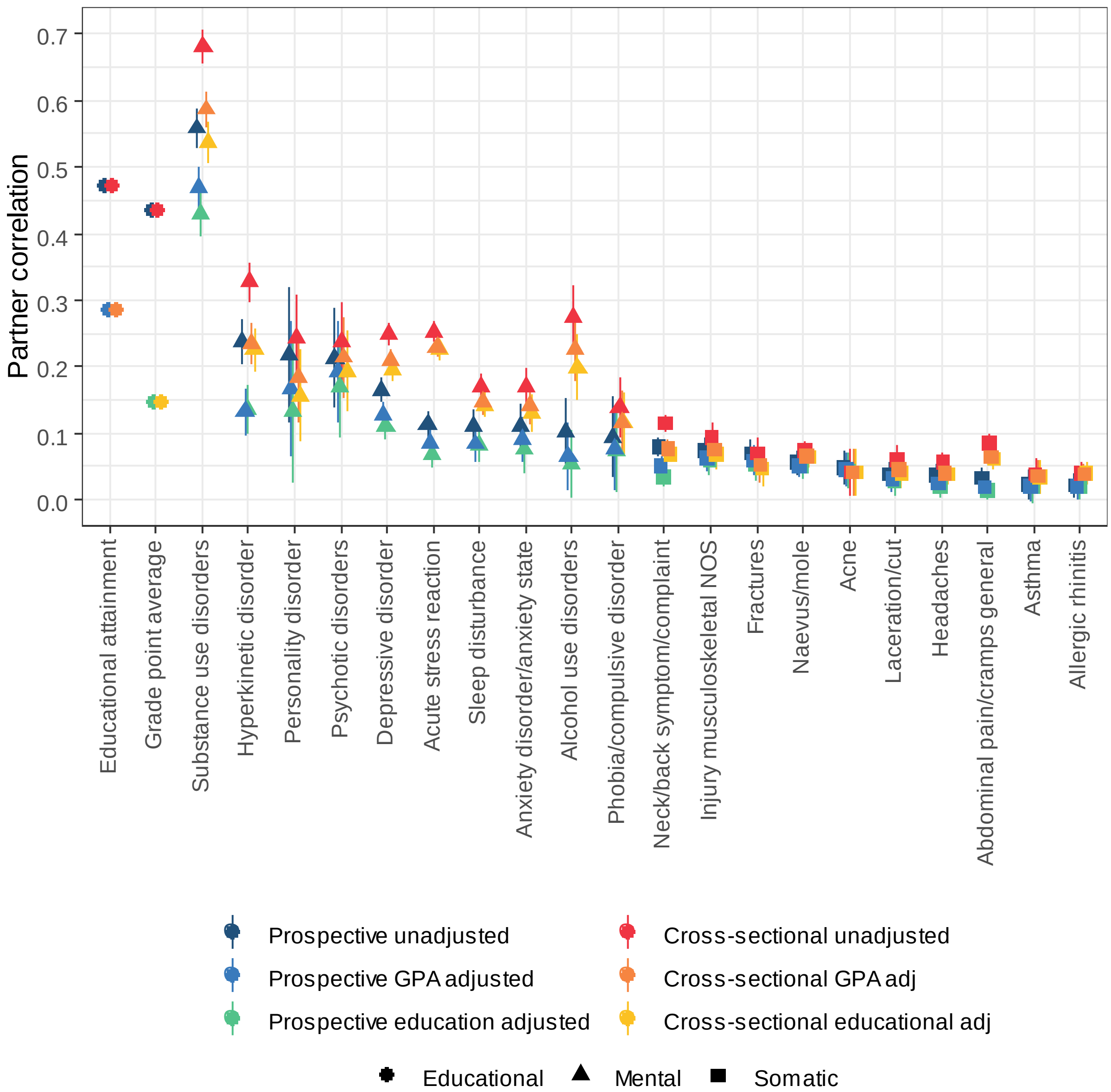


**Supplemental Figure S5**. Correlations between female and male partners for educational outcomes and 10 mental health and 10 somatic health phenotypes 10 to 5 years before they had their first child and cross-sectionally in 2015-2019. Including adjustment for educational attainment and grade point average in both assessments.

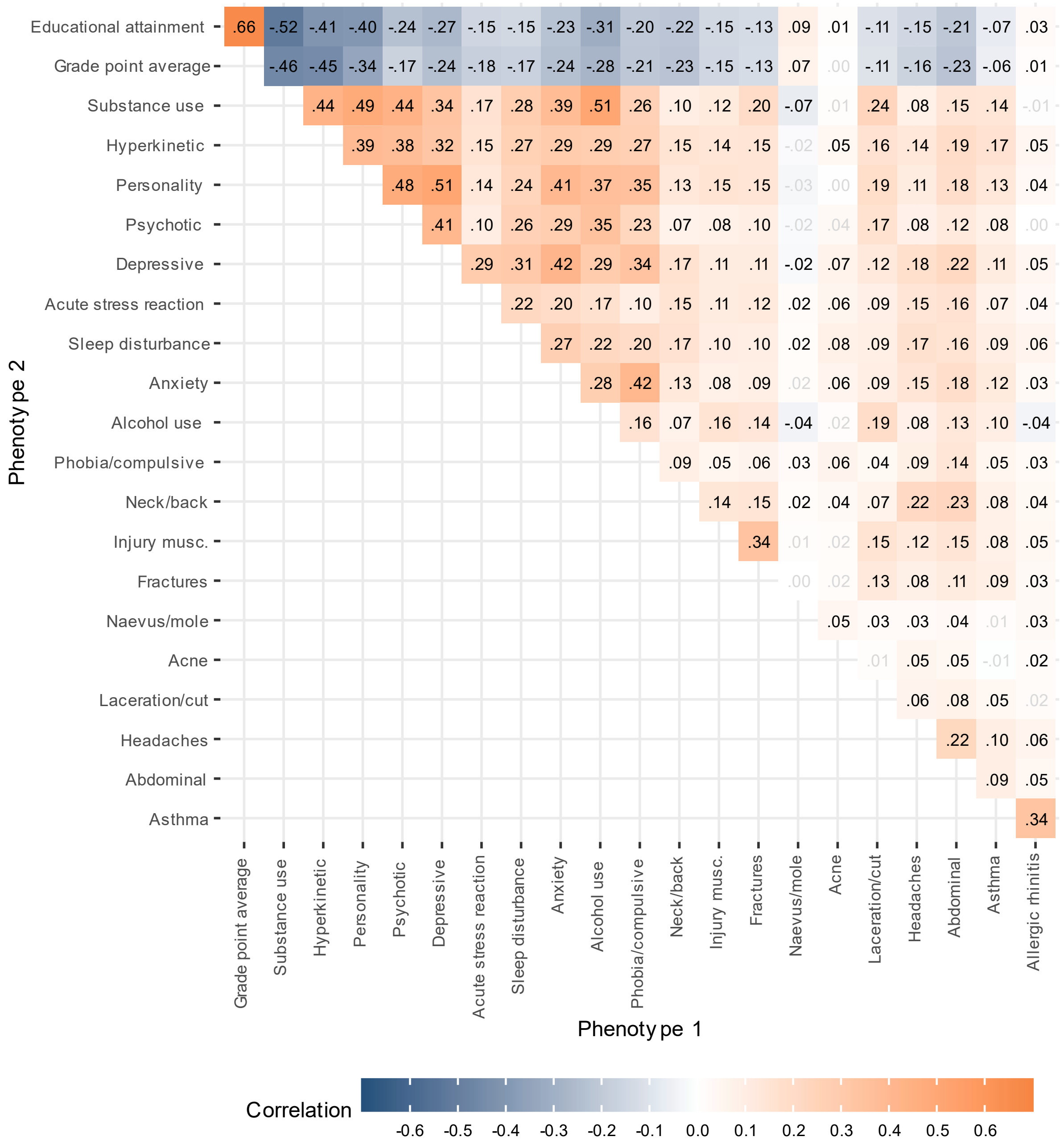


**Supplemental Figure S6.** Within person correlations for educational outcomes, 10 mental health conditions, and 10 somatic health conditions, measured cross sectionally in 2015-2019. Adjusted for age and sex. Correlations shown in black have p-values < 0.05 after adjusting for the false discovery rate.

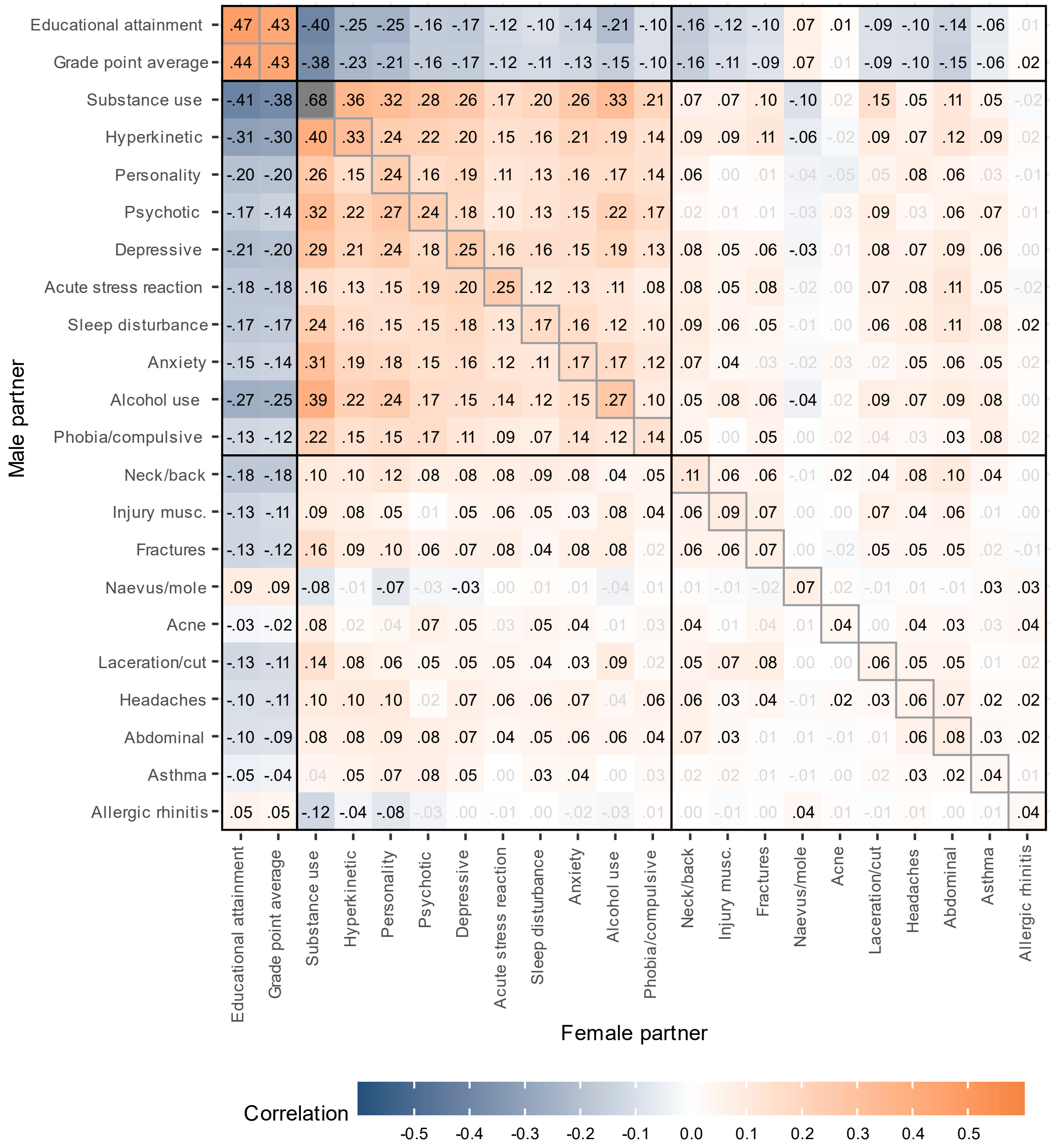


**Supplemental Figure S7.** Within and across-trait partner correlations for educational outcomes, 10 mental health conditions, and 10 somatic health conditions, measured cross sectionally in 2015-2019. Adjusted for age. Correlations shown in black have p-values < 0.05 after adjusting for the false discovery rate.

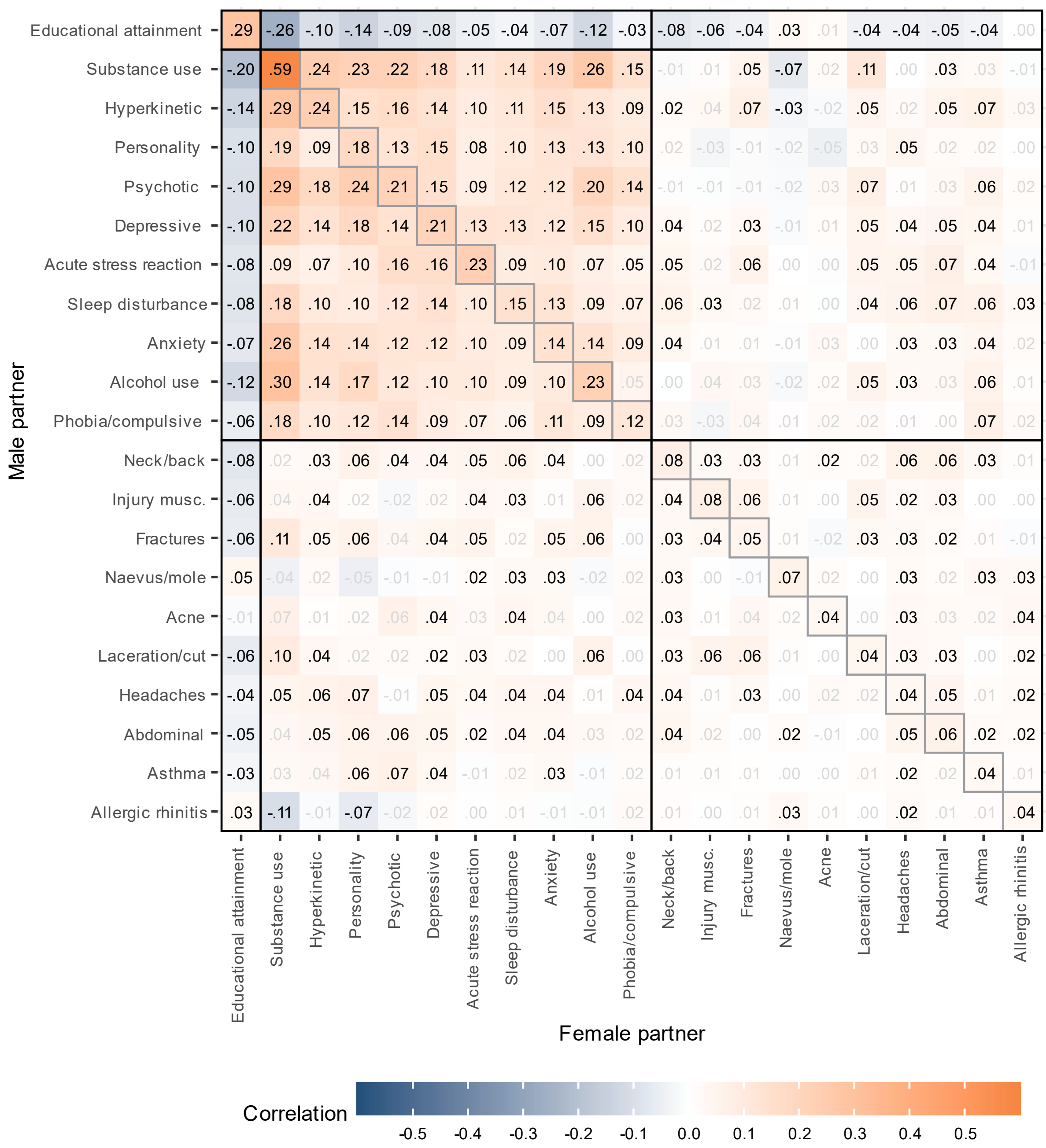


**Supplemental Figure S8.** Within and across-trait partner correlations for 10 mental health conditions, and 10 somatic health conditions, measured cross sectionally in 2015-2019. Adjusted for age and grade point average attainment. Correlations shown in black have p-values < 0.05 after adjusting for the false discovery rate.

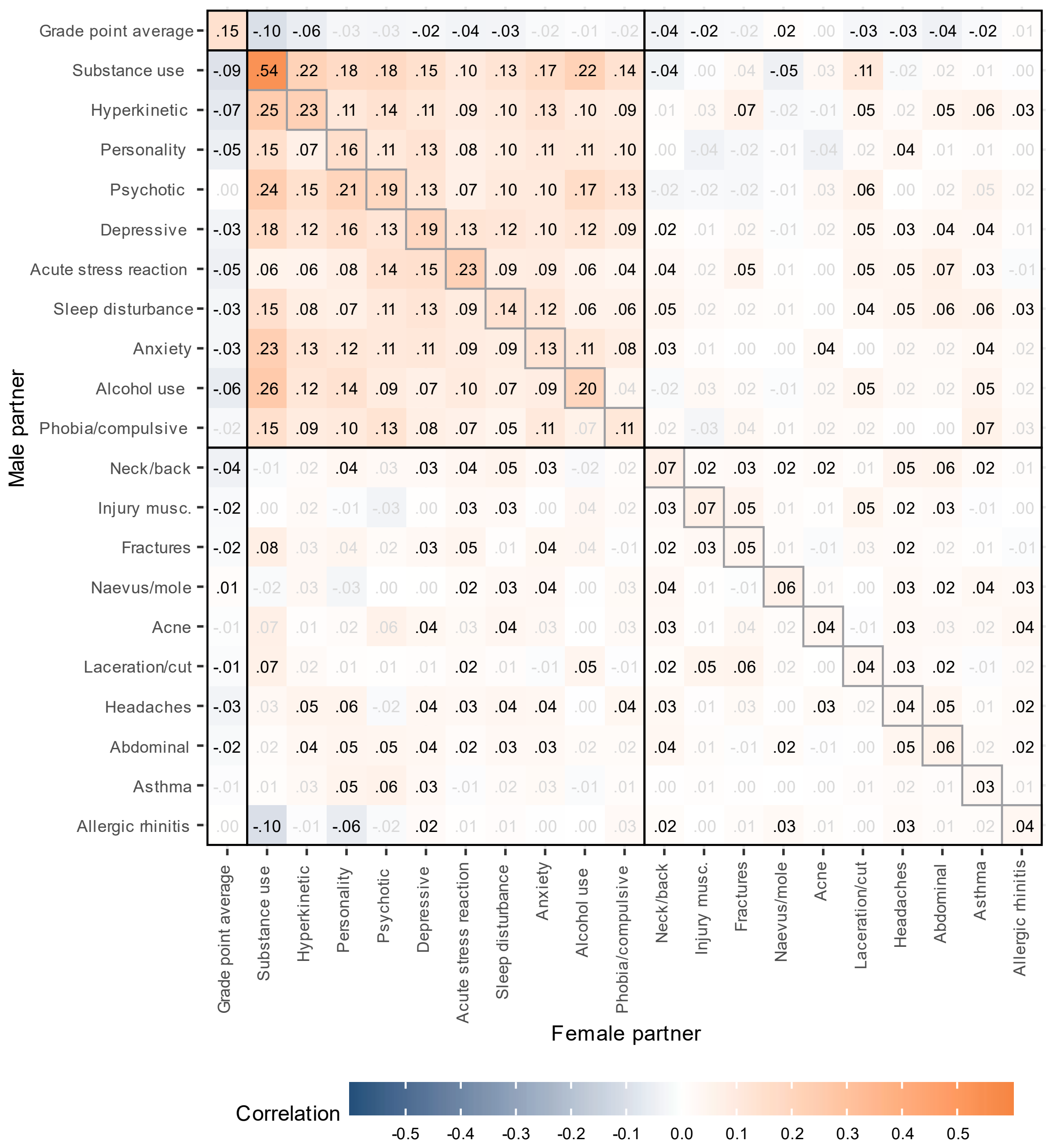


**Supplemental Figure S9.** Within and across-trait partner correlations for 10 mental health conditions, and 10 somatic health conditions, measured cross sectionally in 2015-2019. Adjusted for age and educational attainment. Correlations shown in black have p-values < 0.05 after adjusting for the false discovery rate.

```
library(tidyverse)
N = 10000
IIF = function(m,a,r,x,n,q,e) {
  rpartner = m*a*a+m*x*x+2*x*a+n*n+q*q
  rsibling = a*a*r+e+q*q
  rinlaw = (a*m+x)*r*a+q*q
  rinlaw/(rpartner*rsibling)
}

d = tibble(
  m = runif(N),
  a = runif(N),
  r = runif(N),
  x = 0,
  n = 0,
  q = 0,
  re = runif(N),
  e = re*(1-a^2),
  iif = IIF(m,a,r,x,n,q,e)
)
d %>% filter(iif < 1) %>% count(re > r)
```

**Supplemental Script S1.** R script illustrating of how to simulate the in-law inflation factor (IIF) under various forms of assortment. The current scenario demonstrates indirect assortment and that IIF is above 1.00 except in the implausible cases where $r_e < r_s$.

```
library(polycor)

# dichotomized normally distributed variables
n = 10000
vc = rnorm(n)
va = vc+rnorm(n)
vb = vc+rnorm(n)

va_dich = ifelse(va<2,0,1)
vb_dich = ifelse(vb<2,0,1)

# pearson correlation between original variables
cor(va,vb)

# pearson correlation between dichotomized variables (underestimated)
cor(va_dich, vb_dich)

# polychoric correlation between dichotomized variables (correct)
polychor(va_dich,vb_dich)


# dichotomized skewed variables
n = 100000
vc = rnorm(n)
va = (vc+rnorm(n))^2
vb = (vc+rnorm(n))^2

hist(va)

va_dich = ifelse(va<5,0,1)
vb_dich = ifelse(vb<5,0,1)

# pearson correlation between original variables
cor(va,vb)

# pearson correlation between dichotomized variables
cor(va_dich, vb_dich)

# polychoric correlation between dichotomized variables (overestimated)
polychor(va_dich,vb_dich)
```

**Supplemental Script S2.** R script estimating tetrachoric correlations for variables with underlying normal or skewed distributions. Tetrachoric correlations get overestimated when based on non-normal variables. Written for R.

```
library(tidyverse)
library(polycor)

(randvar=runif(1))

# normally distributed variables

n = 100000
common1 = rnorm(n)
person_i   = common1 + rnorm(n)*randvar      # index
person_ip  = common1 + rnorm(n)*randvar      # index' partner
person_ips = person_ip/2 + rnorm(n)*randvar  # index' partner's sibling
(inlaw)

# correlation matrix
cor(tibble(person_i, person_ip, person_ips))

# observed correlations between indirectly associated variables (e.g.
between in-laws)
cor(person_i, person_ips)

# observed product of correlations (matches the expectation)
cor(person_i, person_ip)*cor(person_ip, person_ips)

# skewed variables
person_i2 = person_i^2
person_ip2 = person_ip^2
person_ips2 = person_ips^2

# correlation matrix
cor(tibble(person_i2, person_ip2, person_ips2))

# observed correlations between indirectly associated variables (e.g.
between in-laws)
cor(person_i2, person_ips2)

# observed product of correlations (still matches the expectation)
cor(person_i2, person_ip2)*cor(person_ip2, person_ips2)

# dichotomized skewed variables
threshold=3
person_i2_dich = ifelse(person_i2<threshold,0,1)
person_ip2_dich = ifelse(person_ip2<threshold,0,1)
person_ips2_dich = ifelse(person_ips2<threshold,0,1)

# correlation matrix
cor(tibble(person_i2_dich, person_ip2_dich, person_ips2_dich))

# observed pearson correlations between indirectly associated variables
(e.g. between in-laws)
cor(person_i2_dich, person_ips2_dich)

# observed product of pearson correlations (lower than the expectation)
cor(person_i2_dich, person_ip2_dich)*cor(person_ip2_dich, person_ips2_dich)

# observed polychoric correlations between indirectly associated variables
(e.g. between in-laws)
polychor(person_i2_dich, person_ips2_dich)

# observed product of polychoric correlations (matches the expectation
again)
polychor(person_i2_dich, person_ip2_dich)*polychor(person_ip2_dich,
person_ips2_dich)
```

**Supplemental Script S3.** R script estimating the product of tetrachoric correlations for skewed variables. This script shows that if the product of two Pearson correlations matches with a third correlation, this is also the case for tetrachoric correlations after dichotomization, even if the underlying variables are skewed. Written for R.
